# Impact of cross-coronavirus immunity in post-acute sequelae of COVID-19

**DOI:** 10.1101/2022.09.25.22280335

**Authors:** Jonathan D. Herman, Caroline Atyeo, Yonatan Zur, Claire E. Cook, Naomi J. Patel, Kathleen M. Vanni, Emily N. Kowalski, Grace Qian, Nancy A. Shadick, Douglas Laffenburger, Zachary S. Wallace, Jeffrey A. Sparks, Galit Alter

## Abstract

Beyond the unpredictable acute illness caused by SARS-CoV-2, one-fifth of infections unpredictably result in long-term persistence of symptoms despite the apparent clearance of infection. Insights into the mechanisms that underlie post-acute sequelae of COVID-19 (PASC) will be critical for the prevention and clinical management of long-term complications of COVID-19. Several hypotheses have been proposed that may account for the development of PASC, including persistence of virus or the dysregulation of immunity. Among the immunological changes noted in PASC, alterations in humoral immunity have been observed in some patient subsets. To begin to determine whether SARS-CoV-2 or other pathogen specific humoral immune responses evolve uniquely in PASC, we performed comprehensive antibody profiling against SARS-CoV-2 and a panel of endemic pathogens or routine vaccine antigens using Systems Serology in a cohort of patients with pre-existing rheumatic disease who either developed or did not develop PASC. A distinct humoral immune response was observed in individuals with PASC. Specifically, individuals with PASC harbored less inflamed and weaker Fcγ receptor binding anti-SARS-CoV-2 antibodies and a significantly expanded and more inflamed antibody response against endemic Coronavirus OC43. Individuals with PASC, further, generated more avid IgM responses and developed an expanded inflammatory OC43 S2-specific Fc-receptor binding response, linked to cross reactivity across SARS-CoV-2 and common coronaviruses. These findings implicate previous common Coronavirus imprinting as a marker for the development of PASC.

**One Sentence Summary:** Through high dimensional humoral immune profiling we uncovered the potential importance of previous common Coronavirus imprinting as a novel marker and potential mechanism of an endotype of PASC.

## Introduction

Since its emergence at the end of 2019, SARS-CoV-2 has caused hundreds of millions of infections across the globe and millions of deaths. SARS-CoV-2 infection causes a broad array of clinical phenotypes ranging from asymptomatic to life-threatening respiratory disease (*1, 2*). While a fraction of individuals succumb to acute infection, the majority of infections resolve within days or weeks (*3-5*). However, emerging data suggest that about 10-50% of previously SARS-CoV-2 infected individuals experience prolonged symptoms that can cause long-term morbidity and compromised life quality (*6, 7*). Post-acute sequelae of COVID-19 (PASC), or “long COVID,” is a general term that encompasses a wide range of symptoms that persist for at least four weeks after initial SARS-CoV-2 infection (*8*). These symptoms predominately present as dyspnea, fatigue, and decreased cognitive function, but other symptoms include loss of olfaction, body aches, dyspnea, congestion, chest pain, fever, and others (*6, 9, 10*). Several risk factors for the development of PASC have been suggested including female gender, pre-existing autoimmune conditions, type 2 diabetes, asthma, and severe COVID-19 (*11, 12*), although none of these risk factors explain the development of symptoms. Thus, the precise causes of PASC as well as the mechanisms that may perpetuate this disease phenotype remain incompletely understood.

Several studies have now begun to probe clinical and immunological perturbations among individuals with PASC compared to non-PASC convalescent controls. Commonly used clinical biomarkers of inflammation including ESR, CRP, D-dimer, as well as organ-specific tissue damage biomarkers (pro-BNP, troponin, neurofilament light chain, glomerular filtration rate) have failed to differentiate between PASC and non-PASC convalescent individuals (*13*). Emerging work has identified increases in total IgM antibody levels, EBV reactivation, and CD8+ T cells against CMV in individuals with PASC (*14*). However, how these markers and risk factors lead to the development of PASC is not well understood. Moreover, PASC likely represents a heterogenous syndrome, with many distinct endotypes, each with distinct pathophysiologic mechanisms. Thus, the identification of biomarkers able to define PASC-within specific endotypes may provide critical insights into the distinct mechanisms and treatment opportunities for management of these long-term complications of COVID-19.

Among potential biomarkers, antibodies represent critical markers of response to vaccination but may also act as indirect markers of historical and current infections(*15*). Thus, changes in pathogen-specific isotype, subclass, and Fc-receptor binding profiles may mark the recency of exposure or immune amplification. Here we deeply explored alterations in the humoral immune response to SARS-CoV-2, additional herpesviruses, common coronaviruses, and childhood vaccines to begin to define whether pathogen-specific antibodies may provide additional insights on PASC. Moreover, given the spectrum of PASC endotypes, here we focused on a single endotype of PASC, applying systems serology (*16*) to a cohort of rheumatic patients with pre-existing rheumatic disease who had experienced mild to moderate COVID-19, half of which developed PASC. We observed a unique humoral immune profile in individuals with PASC compared to matched COVID-19 convalescent individuals without PASC. Patients with PASC had decreased SARS-CoV-2-specific antibody titers and Fc-receptor (FcR) binding capacity but increased humoral responses to endemic coronavirus OC43. Moreover, individuals with PASC had higher avidity IgM antibodies and OC43 S2-focused antibody responses that correlated with their SARS-CoV-2 Spike antibody response, pointing to a potential back-boosted pre-existing OC43 response that may have blunted a de novo antibody response against SARS-CoV-2. In addition, OC43 Spike-specific antibodies were more inflamed, with enhanced FcR binding capabilities and neutrophil activating capacity, in individuals with PASC, marking a recent expansion of OC43 specific immunity. These results point to a potential role for imprinting, or “original antigenic sin,” in the incomplete maturation of the SARS-CoV-2 specific humoral immunity as a marker and potential mechanism in the persistence of symptoms in PASC.

## Results

### PASC Individuals have Lower SARS-CoV-2 Spike immunity

In order to enrich for a specific inflammatory-driven endotype of PASC, we studied humoral immune responses of patients recovered from COVID-19 with pre-existing rheumatic disease. To first determine whether people with PASC developed altered SARS-CoV-2-specific humoral immune responses that could point to differences in viral clearance, we next measured isotype, subclass, and Fc receptor (FcR)-binding profiles of SARS-CoV-2 specific humoral immune responses across the 2 groups of individuals with pre-existing rheumatic disease (Figure 1). Specifically, we measured antibody responses against the SARS-CoV-2 Nucleocapsid and Spike antigens (Figure 1A, B, C, D) and observed significantly lower Spike-specific IgG2 and IgM responses in PASC compared to individuals who did not develop PASC. However, no difference was observed in Spike IgG1 responses or overall Nucleocapsid-specific responses across the 2 groups.

**Figure 1:**
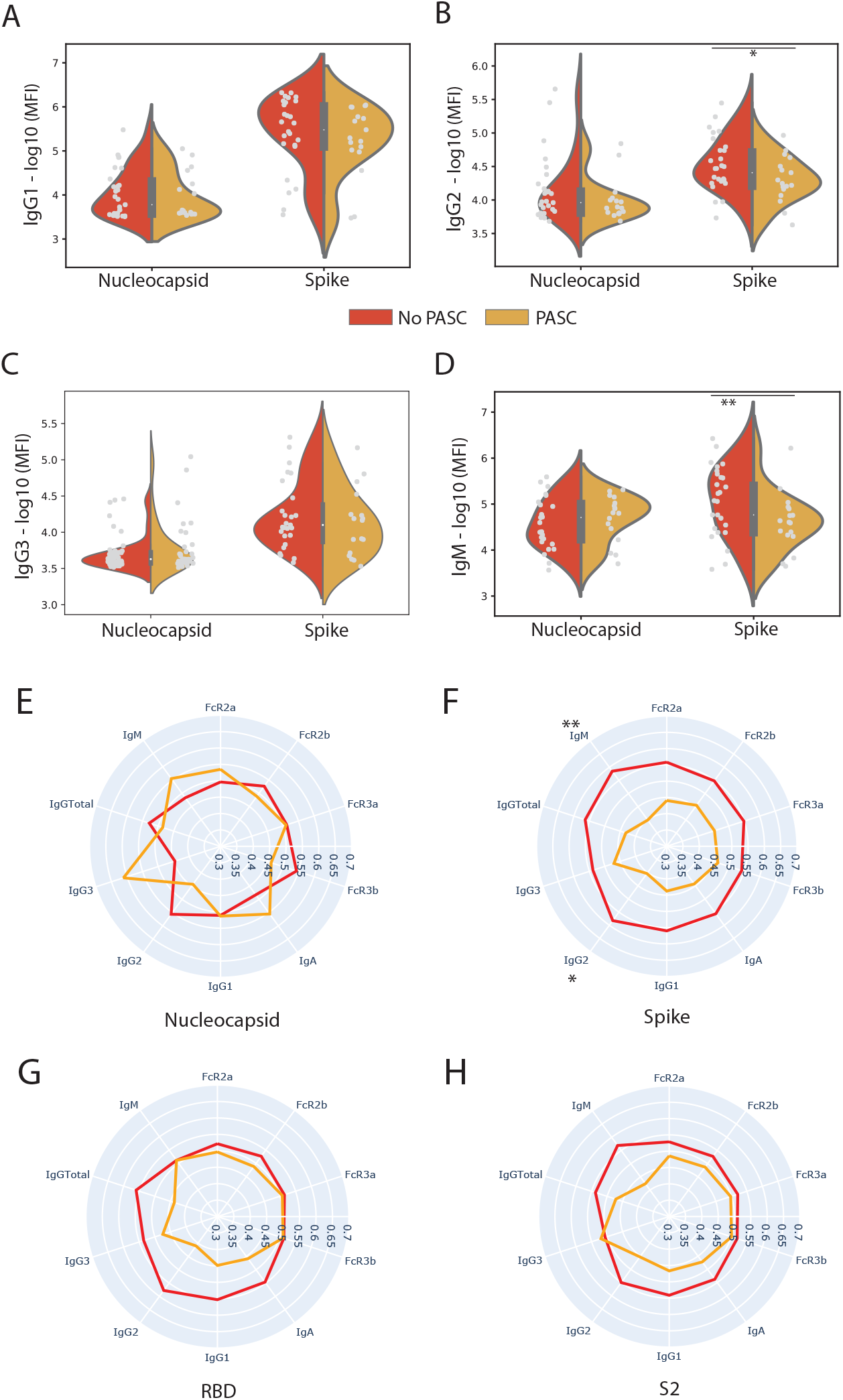
SARS-CoV-2 Spike directed responses are lower in individuals who experienced. (A, B, C, D) The violin plots show the IgG1, IgG2, IgA1, and IgM titer against SARS-CoV-2 Nucleocapsid (N) and Spike, in individuals who experienced PASC (yellow) and those who did not (red). Significance was determined by a two-sided Mann-Whitney U test. (E, F, G, H) The radar plots show the mean percentile rank of antibody titers and Fc receptor (FcR)-binding against Nucleocapsid, Spike (Full Spike, RBD and S2 domain) and for individuals who experienced PASC (Yellow) and those who did not (Red). Significance was determined by a two-sided Mann-Whitney U test. * p = 0.05, p < ** 0.05

To further explore additional differences in the SARS-CoV-2 responses, we measured subclass, isotypes, and Fcγ-receptor binding levels to the Nucleocapsid, Spike, and Spike subdomains RBD and S2 (Figure 1E-H. Nucleocapsid-specific antibody profiles were similar in both groups with an expanded IgG2 and Fcγ3b-binding response in non PASC individuals, and an expanded IgM- and IgG3-response to Nucleocapsid in individuals with PASC. Interestingly, nearly all Spike-specific antibody measurements showed a tendency towards higher levels in individuals without PASC compared to those with PASC, with a significant expansion of IgM-specific Spike responses in individuals without PASC (Figure 1E). A more selective expansion of RBD-specific humoral immune responses was noted between the groups, marked by a tendency towards higher isotype/subclass levels, but not FcR binding levels, in non PASC individuals (Figure 1G). Finally, S2-specific responses displayed a fourth distinct profile, marked by a trend towards higher IgM, IgG1, IgG2, and IgA responses, and specific opsonophagocytic FcRs (FcγR2a and FcγR2b) in individuals without PASC. These data suggest a potential overall blunting of SARS-CoV-2 Spike-specific immunity in individuals with rheumatic disease who develop PASC.

### Distinct multivariate profiles in PASC Reveal OC43 FcγR Binding as a Marker of PASC

Given the trend towards an overall compromised SARS-CoV-2 specific humoral immune response among individuals with PASC, we next sought to determine whether other changes in the humoral immune response, to common vaccines or common human pathogens, could provide additional insights into differences in humoral immune responses in SARDS individuals with or without PASC. Thus, we profiled antibody isotype titers and FcR-binding levels across an array of antigens, including common vaccine antigens (Tetanus, Rubella, Measles, Mumps), herpesviruses (herpes simplex 1 - HSV-1, cytomegalovirus -CMV, Epstein Barr virus -EBV, Varicella Zoster virus - VZV), other coronaviruses (SARS-CoV-1 Spike/S1/S2, OC43 Spike/S1/S2, HKU1 Spike/S1/S2) and other human-infecting pathogens (Influenza, Respiratory Syncytial virus (RSV), and Staphylococcus aureus) and control pathogens (Ebola). A minimal set of multivariate differences in antibody profiles were identified using elastic net regularization followed by a partial least squares discriminant analysis (PLSDA) to visualize differences across the groups (Figure 2A). Multivariate analysis of this broader humoral immune response across PASC and non-PASC individuals revealed significantly different overall antibody profiles across the 2 groups to common pathogens and vaccine antigens. The model was validated with a 5-fold cross validation framework and performed significantly better than null models (Supplementary Figure 1A). As few as 12 of the total 270 captured antibody features were selected by elastic net regularization to drive this multivariate profile difference, marked by the enrichment of 7 features in the non-PASC individuals and 5 features in the PASC individuals (Figure 2B). Higher levels of RSV-specific IgM/IgG3, Influenza HA-specific FcγR2a/2b binding levels, SARS-CoV-1 S1-specific IgA, and Tetanus specific IgG/IgM were all selectively enriched among non-PASC individuals (Figure 2B, F, G, H). Conversely, CMV gB-specific FcγR2a/3b/IgM and OC43 Spike-specific FcγR3a/3b binding levels were selectively enriched among individuals with PASC (Figure 2B), all of which were significant at a univariate level (Figure 2C-E). Overall, the data suggested that PASC was associated with a significant alteration in the overall polyclonal humoral immune response to common pathogens and vaccines.

**Figure 2:**
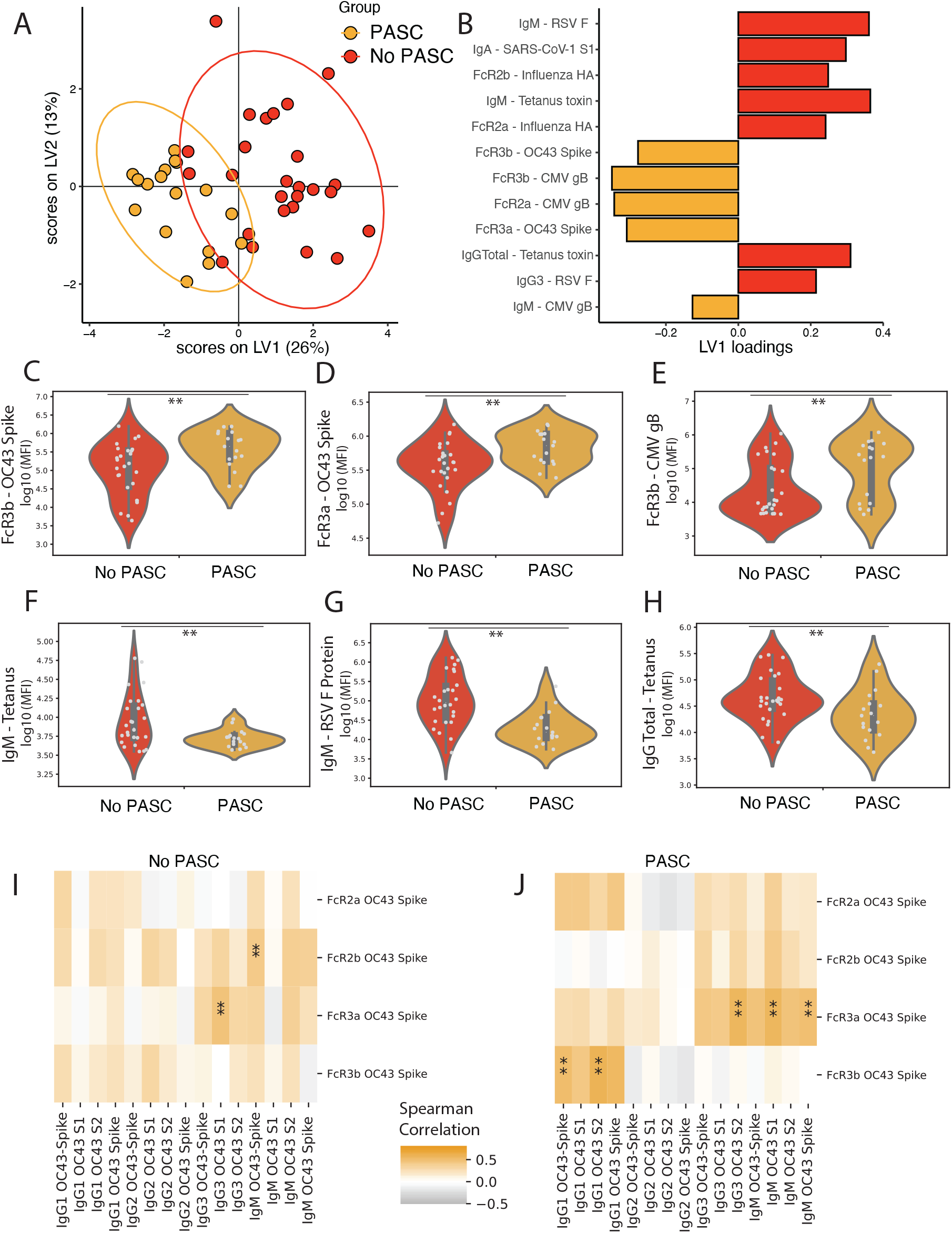
Differences in non-SARS-CoV-2 humoral response in individuals who experienced PASC and those who did not. (A) A partial least squares discriminant analysis (PLSDA) was built using Elastic Net selected features. Each dot represents a sample, and the color represents the group. The ellipses show the 95% confidence interval. (B) The bar plot shows the loadings along latent variable 1 (LV1) for the Elastic Net selected features used to build the PLSDA in (A). The color indicates the group that the feature was enriched in. (C - H) The violin plots show the top three Elastic Net features enriched in the (C, D, E) PASC group (OC43 Spike FcR3b, OC43 Spike FcR3a, and CMV FcR3b) and (F, G, H) no PASC group (Tetanus IgM, Tetanus Total IgG, and RSV IgM). Significance was determined by a two-sided Mann-Whitney test, ** p < 0.05. (I, J) The two heatmaps shows the correlation of OC43 Spike Fc-receptor binding and OC43 isotypes and Isotypes in (I) PASC and (J) No PASC individuals. Positive and negative correlation are represented respectively by orange and gray colors. ** indicates a p-value < 0.05.

Since prior exposure to CMV occurs in approximately 50% of United States residents(*17*), we re-evaluated the selected CMV features for serostatus.(*17*) When we selected only seropositive individuals, defined by CMV-specific IgG levels, we did not find significant differences in CMV gB FcγR2a/3b/IgM (Supplementary Figure 1B, C), suggesting that the quality or type of CMV antibodies did not differ between individuals who did and did not experience PASC, but that rather potential imbalances in CMV-prevalence between both groups may skew the importance of these antibody features in our model. Rather, CMV IgG and FcγR weakly correlated with most of the array of antigens tested (Supplementary Figure 1D). This argues against reactivation of CMV as a key marker of PASC in this cohort.

Given the unexpected signature of expanded OC43 immunity in individuals with PASC, and the potential role of OC43 imprinting in original antigenic sin,(*18-23*) we next aimed to explore the coordination of the OC43-specific humoral immune responses with the SARS-CoV-2 response across the groups. Spearman correlations of OC43 FcγR-binding features (selected by the model) and other OC43 titers in both groups revealed different overall antibody architectures across the groups (Figure 2I, J). While individuals who did not develop PASC exhibited a general coordination across all OC43-specific Fc-receptor binding and isotype/subclass responses (Figure 2I), individuals with PASC showed 2 patterns (Figure 2J). First, OC43-specific IgG1 responses were strongly correlated with neutrophil activating OC43-specific FcγR3b binding antibodies, but not other OC43-specific subclasses and isotypes. The second profile included OC43-specific FcγR2a, FcγR2b, and FcγR3a binding antibodies that were correlated with OC43-specfic IgG3 and IgM responses, isotypes classically induced during an acute response following the recent selection and activation of B cells(*24*). These data point to an overall shift in the OC43-specific humoral immunity in individuals with SARD who develop PASC.

### Patients with PASC Have High Avidity, Matured OC43 S2 responses

To further dissect the unique inflamed OC43-specific humoral immune response in subjects with PASC, we next plotted the mean percentile rank of the OC43 Spike isotypes, subclass, and FcγR-binding across the two groups (Figure 3A). OC43 Spike FcγR-receptor binding antibodies were significantly elevated in individuals with PASC while IgG3 and IgM titers were not increased, pointing to the expansion of highly inflammatory IgG1 responses rather than the evolution of new OC43 antibodies in individuals with PASC, that would transition through IgM/IgG3(*25*).

**Figure 3:**
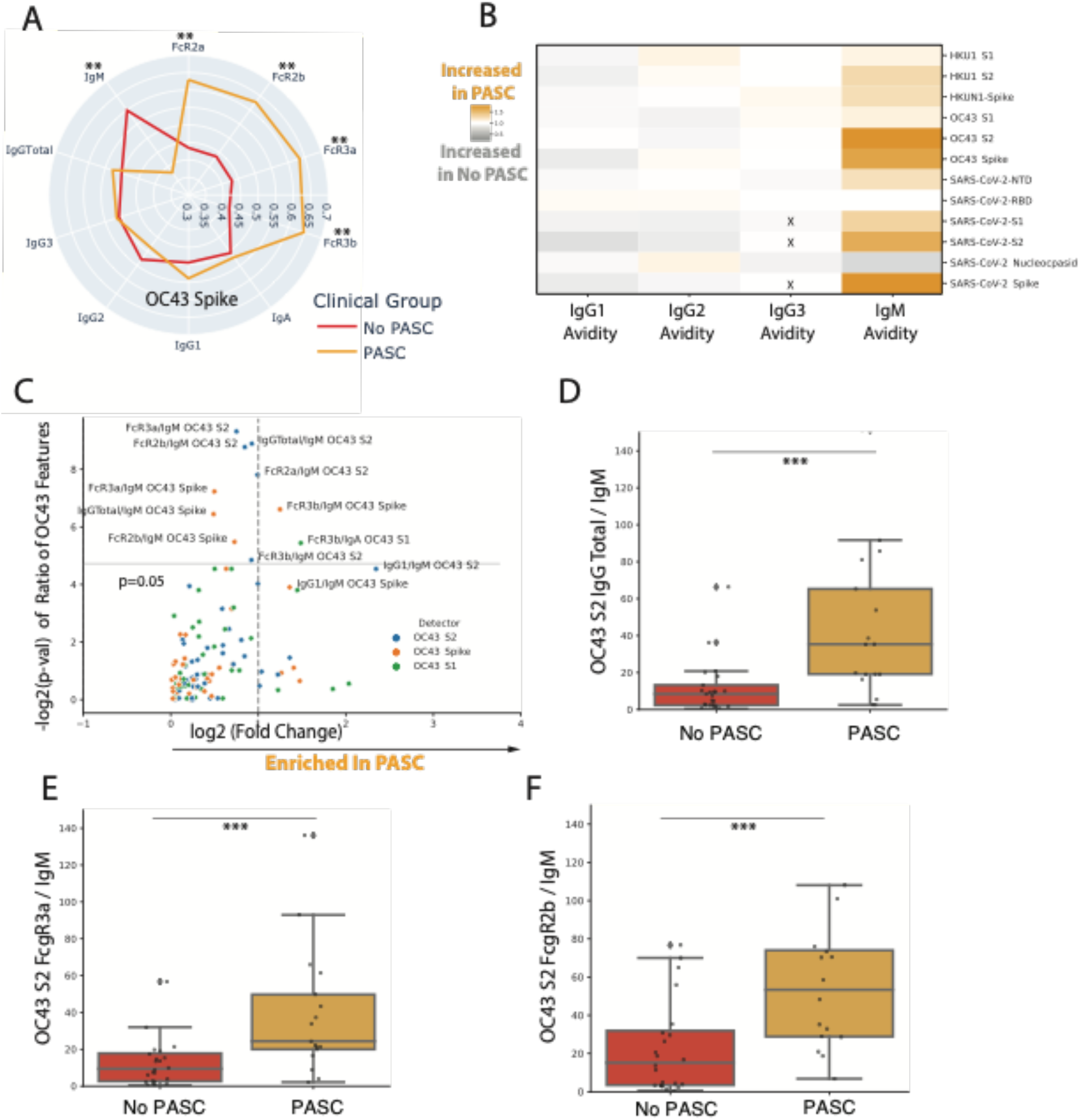
Patients with PASC Have High Affinity Matured their OC43 S2 responses. (A) The radar plots show the normalized values for antibody titers and Fc receptor (FcR)-binding against OC43 Spike for individuals who experienced PASC (Yellow) and those who did not (Red). Significance was determined by a two-sided Mann-Whitney U test. p < ** 0.05. (B)The heatmap shows the ratio of Avidity in PASC over No PASC individuals. Avidity of IgG1, IgG2, IgG3, and IgM was measured for SARS CoV-2, OC43, and HKU1 Spike, S1 domain, and S2 domain antigens. Orange represents avidity measurements increased in individuals with PASC and grey represents avidity measurements increased in patients without PASC. X indicates this value was not measured. (C) Volcano plot of Antibody Measurement ratios for OC43, OC43 S1, and OC43 S2 antigen measurements. All ratio permutations of isotype, subclass, and Fc-receptor binding measurements are included and significance between PASC and No PASC individuals was tested with two-sided Mann Whitney U-testing. Ratios with a log 2-Fold change (x-axis) are included in the plot. Log2 fold change with negative values suppressed (x-axis) and –log10 p-value (y-axis) are plotted and points are colored by their antigen. (D, E, F) Box plots show selected ratios in PASC (Yellow) and No PASC (Red) individuals for (D) the ratio of IgGtotal/ IgM for OC43 S2 domain, (E) of FcR3a/IgM for OC43 S2 domain, and (F) of FcR2b/IgM for OC43 S2 domain. Significance was determined by a two-sided Mann-Whitney U test. *** p< 0.005.

To further probe whether the selective expansion of OC43-specific responses was related to the evolution of new or recall of pre-existing immune responses, we next probed the avidity of the SARS-CoV-2 and common Coronavirus humoral response (Figure 3B). IgG1, IgG2, and IgG3 responses did not exhibit enhanced avidity among individuals with PASC compared to individuals who did not have PASC. However, despite having lower OC43 IgM levels (Figure 3A), individuals with PASC possessed more avid IgM-responses to the more highly conserved S2-domain of both OC43 and SARS-CoV-2 (Figure 3B) pointing to the selective affinity maturation of IgM responses across coronaviruses. To further probe the relationship of this unexpected IgM response to OC43 S2 in PASC, we compared all antibody features to IgM avidity by calculating ratios (example IgM/IgG3, IgM/IgA, etc.) of all measured antibody features for each OC43 antigen (Spike, S1, and S2) in individuals with and without PASC (Figure 3C). PASC was associated with a significant selective induction of highly functional OC43 S2-specific Fc-receptor binding responses in relation to OC43 specific IgM levels (Figure 3 C). Moreover, at a univariate level, individuals with PASC clearly experienced a selective expansion of OC43 S2-specific IgG and FcγR specific responses at the expense of a loss of IgM responses (Figure 3E-G) but not of OC43 S1 (Supplementary Figure 2A, B), pointing to a unique evolution of the humoral immune response among individuals with PASC. Further, this unique pattern was also present in SARS-CoV-2 S2 responses, but to a lesser extent, suggesting that the OC43 humoral response may have imprinted and influenced the evolution of the SARS-CoV-2 response in participants with PASC (Supplementary Figure 2 C, D). Thus, these data suggest that individuals with PASC experience a unique selective expansion of more inflammatory FcγR binding IgG to the OC43 S2 in contrast to the individuals who do not have PASC.

### Individuals with PASC Have a Skewed OC43 and SARS-CoV-2 S2 Inflammatory Antibody Response

To examine the relationship between the OC43- and SARS-CoV-2-specific immunity, we first looked at the Spearman correlation between OC43 Spike IgM, that was uniquely regulated in PASC (Figure 3A, D), and SARS-CoV-2 FcγR binding responses. While limited relationships were observed in individuals who did not develop PASC between OC43-specific IgM responses and the overall response to SARS-CoV-2, OC43 IgM levels were positively correlated with all measured SARS-CoV-2 S2 FcγRs in individuals with PASC, with a highly significant relationship between OC43 Spike IgM levels and SARS-CoV-2 S2-specific FcγR binding levels (Figure 4A). This suggested that the high avidity OC43 IgM response in individuals with PASC was highly coordinated with the development of a FcR-binding SARS-CoV-2 S2-specific humoral immune response.

**Figure 4:**
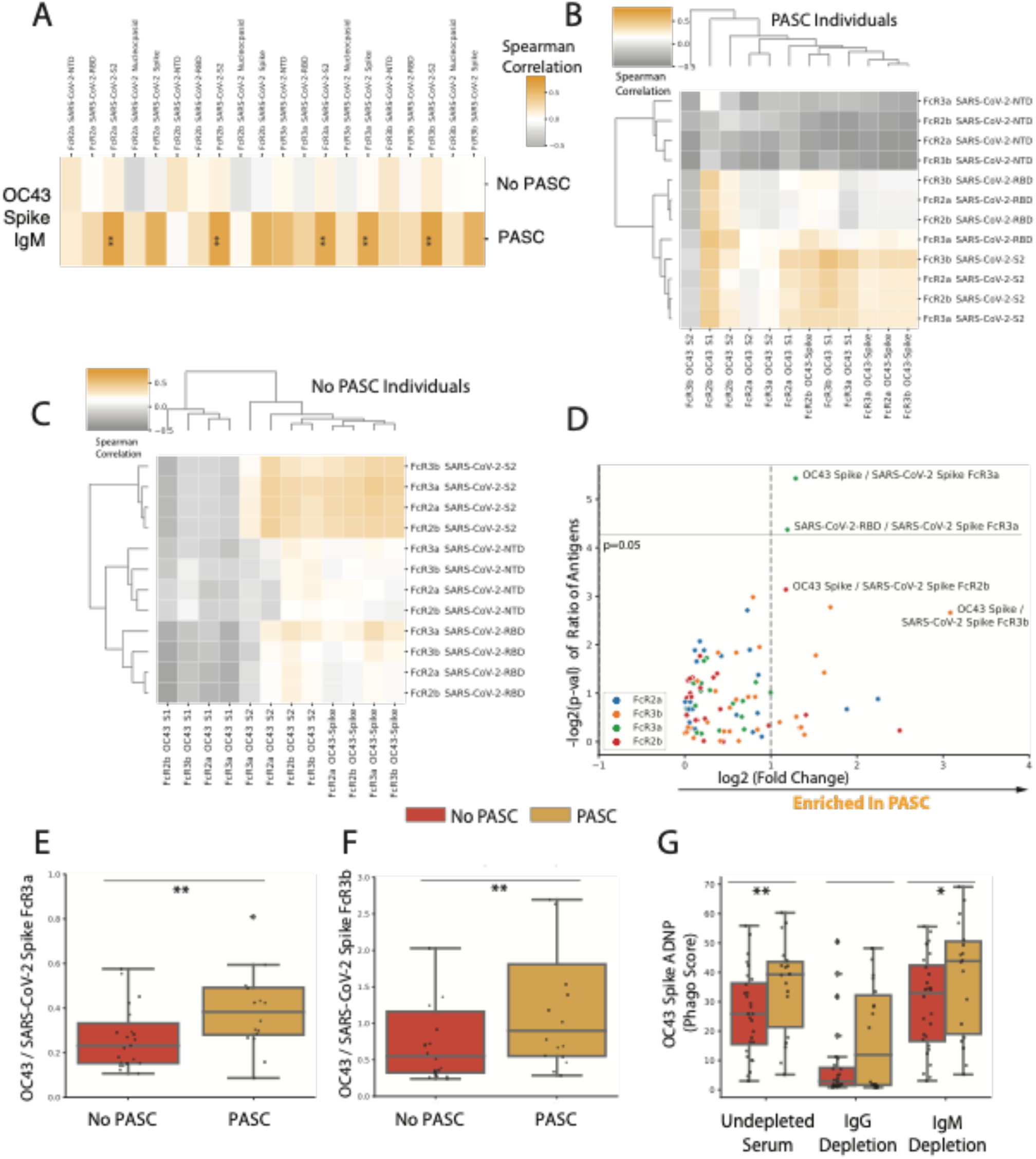
PASC Individuals Have a Stronger OC43 than SARS-CoV-2-S2 Inflammatory Antibody Response. (A) The heatmap shows Spearman correlation of IgM OC43 features with SARS-CoV-2 Fc receptor binding capacity in PASC and No PASC individuals. Positively correlated features are colored orange and negatively correlated features are colored grey. Significance is indicated as follows * p < 0.1, p < ** 0.05, p< 0.005. (B) This clustered heatmap shows Spearman correlation of OC43 FcRs (x-axis) and SARS-CoV-2 Spike subdomains (NTD, RBD, S2) (y-axis) in PASC individuals. Positively correlated features are colored orange and negatively correlated features are colored grey. (C) This clustered heatmap shows Spearman correlation of OC43 FcRs (x-axis) and SARS-CoV-2 Spike subdomains (NTD, RBD, S2) (y-axis) in individuals without PASC. Positively correlated features are colored orange and negatively correlated features are colored grey. (D) The volcano plot shows the Ratio of Fc-receptor binding of OC43 and SARS-CoV-2 antigens measured. All ratio permutations of OC43 and SARS-CoV-2 antigens Fc-receptor binding measurements are included and significance between PASC and No PASC individuals was tested with a two-sided Mann Whitney U-testing. Ratios with a log 2-Fold change (x-axis) are included in the plot. Log2 fold change (x-axis) and –log10 p-value (y-axis) are plotted and points are colored by their antigen. (E, F) Box plots show selected ratios in PASC (Yellow) and No PASC (Red) individuals for (E) the ratio of OC43 Spike/SARS-CoV-2 Spike for FcR3a binding and (F) of OC43 Spike/SARS-CoV-2 Spike for FcR2b binding. (G) The box plots show Antibody-Dependent Phagocytosis of OC43 Spike in PASC (Yellow) and No PASC (Red). Samples were depleted for IgG and IgM (x axis). Significance between PASC and No PASC individuals was tested with two-sided Mann Whitney U-testing. * p = 0.05, ** p < 0.05.

To further define whether this altered coordination of OC43 Spike specific immunity skewed SARS-CoV-2 S2-specific immunity alone, or also impacted the evolution of humoral responses to other domains on the Spike antigen, we next probed the relationship between OC43 Spike FcγR binding responses and domain-specific responses to SARS-CoV-2 (Figure 4B, C). Differences were present in the overall architecture of the OC43/SARS-CoV-2 functional antibody response across PASC (Figure 4B) and non-PASC (Figure 4C), marked by a trend towards overall negative correlations between OC43 FcγR-binding levels and SARS-CoV-2 NTD-specific responses only in individuals with PASC. Moreover, SARS-CoV-2 S2-specific responses were positively correlated with several Spike, S1, and S2-specific OC43-specific FcγR binding responses in individuals with PASC, but these were not all OC43-S2-specific, suggesting that SARS-CoV-2 S2 specific immunity may not have evolved solely from pre-existing OC43 S2-specific responses, but a SARS-CoV-2 S2-specific response may have emerged in a coordinated manner with broad cross-Spike-specific pre-existing OC43 immunity (Figure 4B).

To ultimately understand the relationship between OC43-specific immunity and the evolution of SARS-COV-2 immunity, we compared FcγR-binding levels by calculating ratios of all measured FcγR-binding antibody responses of SARS-CoV-2 and OC43 Spike (full length, RBD, S1, S2) in individuals with and without PASC and then tested which ratios were enriched in PASC individuals (Figure 4D). OC43/SARS-CoV-2 Spike FcγR3a was significantly enriched in individuals with PASC (Figure 4D, E and F). Additionally, OC43/SARS-CoV-2 Spike FcγR2b and FcγR3b binding trended towards also being enriched in PASC (Figure 4C and E). Furthermore, linked to higher FcγR3b binding, individuals with PASC harbored higher levels of OC43-sepcific antibody-dependent neutrophil phagocytosis (Figure 4F); higher neutrophil activity has previously been linked with enhanced inflammation and disease severity(*26*). Moreover, depletion studies pointed to the dominant role of IgG, rather than IgM, as the major driver of this enhanced OC43-specific antibody effector function in PASC (Figure 4G, Supplementary Figure 3B, C). Thus, collectively, these data point to an altered architecture within the SARS-CoV-2 response in individuals with PASC, significantly influenced by OC43-specific immunity. Given the more modest evidence of this relationship within individuals who do not develop PASC, these data point to potential back-boosting of the OC43-specific immunity in PASC individuals, potentially blunting the SARS-CoV-2 response, and resulting in the aberrantly persistent production of inflammatory antibodies that may drive less effective clearance of SARS-CoV-2.

## Discussion

PASC affects a high proportion of individuals who are infected with SARS-CoV-2(*27, 28*). Over 40% of COVID-19-recovered individuals have persistent symptoms 90 days later (*29*) Further in individuals with pre-existing rheumatic disease, up to 45% of COVID-19 recovered individuals have persistent symptoms 28-days later(*30*).(*29*). Preliminary work to identify biomarkers of PASC has identified increased EBV seromarkers or reactivation (*14, 31*), presence of blood viral SARS-CoV-2 RNA (RNAemia) (*32*),, total IgM antibody levels, CD8+ T cells against CMV (*14*), and persistent Spike protein(*33*). Though some of these markers, including EBV reactivation and RNAemia, may be markers of COVID severity of disease, the causal relationship of others with PASC remains unclear. Understanding the mechanisms that lead to the development of PASC is challenging due to its broad syndromic definition likely with multiple endotypes each with distinct pathophysiologic mechanisms. SARS-CoV-2 can induce the production of autoantibodies(*34, 35*), cause exacerbated tissue/organ damage(*2*), lead to neurologic inflammatory complications(*36, 37*), exacerbate pre-existing SARDS symptoms(*30*), and cause the new onset of SARDs (*38, 39*). To focus in on a single endotype, here we used systems serology to profile a cohort of COVID-19 rheumatic patients months after initial COVID infection who were clinically evaluated for PASC. We observed lower SARS-CoV-2 Spike and S2 antibody responses in individuals who experienced PASC on a univariate level. Using multivariate modeling, we observed individuals who experienced PASC had higher CMV- and OC43-directed inflammatory antibodies. While the CMV-specific response was largely driven by an imbalance of CMV-seroprevalence in our cohort, we observed that the OC43 response was driven by high avidity IgM OC43 antibodies and class-switched FcγR binding OC43 responses that were inversely correlated with the quantity and quality of the SARS-CoV-2 response, pointing to a potential role for previous common Coronavirus back-boosting as a driver of incomplete SARS-CoV-2 antibody generation in individuals with SARDS who develop PASC. These results point to immunological imprinting in PASC that may result in the generation of incomplete SARS-CoV-2 immunity, control, and clearance.

OC43, one of the human endemic coronaviruses responsible for the common cold, and SARS-CoV-2 are both beta-coronaviruses, exhibiting high sequence conservation (*40*). Back-boosting, imprinting, or original antigen sin, are immunologic phenomena where the immune system expands pre-existing memory responses at the time of a new infection, however these responses preferentially recognize a closely related pre-existing ancestor of the current infection. In the setting of Influenza infection, immunization with contemporaneous strains often results in the expansion of antibody responses that both preferentially recognize previously encountered strains as well as the new strains. This recall expansion has been proposed to hamper the ability to drive de novo, distinct immune responses with greater breadth (*41*). In the setting of SARS-CoV-2, imprinting has also been observed, whereby both repeated boosting with the same antigen or infection with similar historical strains resulted in the generation of antibody responses with more limited capacity to neutralize divergent variants of SARS-CoV-2(*42*). Importantly, cross-reactive responses to SARS-CoV-2 have been documented in pre-pandemic sera and PBMCs(*43*) and have been linked to milder acute infection (*18, 21, 44, 45*). Moreover, early studies pointed to evidence of back-boosting of humoral responses to seasonal coronaviruses, including OC43 (*23, 44*), and the expansion of functional OC43 responses were linked to milder acute disease(*18*). Conversely, some studies have suggested that antibody responses to seasonal coronaviruses are enriched in severe COVID-19 (*20, 22, 46*), potentially inhibiting the evolution of SARS-CoV-2-neutralizing and non-neutralizing antibody responses(*46, 47*). Likewise, here we have found that back-boosted functional OC43-specific responses were enriched in individuals with rheumatic disease who developed PASC. The observations of affinity matured IgM and persistently inflamed OC43-specific FcγR-binding antibodies in individuals with PASC, in the setting of diminished SARS-CoV-2 immunity long after initial COVID-19 illness, points to potential immunologic imprinting that resulted in blunted maturation of SARS-CoV-2 immunity. Rather, the recall-favored OC43-specific immunity may less effectively clear SARS-CoV-2 infection and lead to persistent inflammation and symptomatology associated with a specific endotype of PASC.

Given the limited evidence of widespread co-circulation of OC43, at the time of the COVID-19 pandemic(*48-50*), it is unlikely that the expanded OC43-specific response observed in individuals with PASC are related to infection with a common Coronavirus. Instead, the data point to a more likely back-boosting event that resulted in an attenuation of the evolution of a SARS-CoV-2 specific immune response. Interestingly, vaccination, aimed at boosting SARS-CoV-2 immunity, was originally proposed as a potential mechanism to improve PASC outcomes. However, vaccination in individuals with PASC has not translated to an amelioration of PASC despite the induction of SARS-CoV-2 specific antibodies(*51, 52*). Whether this is related to permanent imprinting that cannot be rescued with additional boosting, or to incomplete competition of new vaccine induced antibodies with pre-existing OC43-specific responses remains unclear. Instead, emerging data point to a potential persistence of SARS-CoV-2 Spike antigen in individuals with PASC (*33, 53-56*), despite the presence of SARS-CoV-2 antibodies. Moreover, prolonged viral replication and evolution, has also been noted in immunocompromised hosts(*57*). Though viral RNA weeks to months after acute respiratory infection in the gastrointestinal tract has raised the possibility of it as a possible site of prolonged SARS-CoV-2 infection, persistent infection of immunocompetent individuals has yet to be demonstrated via gold standard techniques such as viral culture(*55, 58*). Further, circulating Spike-containing extracellular vesicles were demonstrated in a small cohort of patients with PASC months after infection (*33*). Thus, it is plausible that imprinted immunity in individuals with PASC may lead to incomplete clearance of viral antigens, viral particles, and virus-containing extracellular vesicles that may persist as immune complexes and cause inflammation and disease. However, whether monoclonal antibodies, antivirals, or new imprint-breaching vaccines will be able to clear any remaining virus, antigens, or particles remains to be defined.

The clinical term PASC has been applied to this wide spectrum of disease and likely includes both systemic and organ-specific pathologies. Here we focused on a specific endotype of PASC, observed in individuals with previous rheumatic disease. We observed a unique shift in the inflammatory architecture pointing to the potential role of original antigenic sin in the incomplete maturation of the SARS-CoV-2 humoral immune response. Thus, whether imprinting will also be observed in the setting of other PASC endotypes remains unclear, but points to an immunologic deficiency in this patient population that may be strategically targeted to develop potential PASC therapies. Moreover, markers like OC43 Spike-specific FcγR binding, found in this study, could identify individuals at high risk of developing PASC, and help to identify and enroll individuals in a strategic manner into future clinical prophylactic and therapeutic trials of PASC therapies.

### Limitations to the study

There are several limitations to this study. First, this study only measured antibody measurements months after initial COVID symptoms. Therefore, we were unable to determine if the observed differences between individuals who experienced PASC and those who did not were triggered by initial SARS-CoV-2 infection. Secondly, this cohort consisted only of individuals who have rheumatic disease. Whether these observed differences in antibody features between individuals who do or do not develop PASC in this SARDS cohort is generalizable in other populations is unclear. In addition, due to a low number of samples, we were unable to associate certain antibody features with different symptom profiles. Future studies with a larger numbers of individuals with PASC should aim to understand how differences in immunological response led to diverse PASC symptoms. Still, this study shows for the first time that inflammatory antibody responses to common coronaviruses are associated with PASC symptoms, enhancing our understanding of the mechanisms that cause this debilitating syndrome.

## Methods

### Study Design

We analyzed patients recovered from COVID-19 with pre-existing rheumatic disease due to their propensity for inflammation and autoantibody production, which could make them more vulnerable to PASC and enrich for specific inflammatory-driven endotypes. Since March 1, 2020, all SARD participants with PCR- or antigen-confirmed SARS-CoV-2 were identified in the Massachusetts General Brigham (MGB) HealthCare system, supplemented by referrals from rheumatologists of participants who were diagnosed with COVID-19 outside of the MGB system(*30, 59-63*). Only individuals who did not require hospitalization for COVID-19 were included in our study. As in previous studies(*30, 59-63*), we excluded participants treated solely for osteoarthritis, fibromyalgia, mechanical back pain, gout, or pseudogout without a SARD since these conditions are not typically treated with systemic immunomodulators and are often managed by non-rheumatologists. Thus, all patients included had a confirmed pre-existing SARD diagnosis with laboratory-confirmed outpatient COVID-19. All participants provided informed consent prior to participation in the study and the clinical study was approved by the Mass General Brigham Institutional Review Board.

In this analysis, 17 SARD participants with PASC and 26 without PASC were profiled (Table 1). The majority of patients received a COVID-19 vaccine (95%) and were female (79%). Rheumatoid arthritis was the most common rheumatic disease (35%) followed by psoriatic arthritis (19%); tumor necrosis factor (TNF) inhibitors were the most common therapy used (26%) followed by methotrexate (14%) or hydroxychloroquine (12%) monotherapy.

**Table 1:** Clinical Characteristics of SARD Patient Cohort. Number (percentage) of participants with selected demographic, COVID-19 associated, rheumatic disease, and immune modulator therapy characteristics.

|  | **Overall**<br>**(n=43)** |
| --- | --- |
| **Median age (IQR)** | 56 (46, 66) |
| **Female sex, n (%)** | 34 (79%) |
| **Race, n (%)** |  |
| White | 38 (88%) |
| Black | 3 (7%) |
| Asian | 1 (2%) |
| Other | 1 (2%) |
| **Hispanic, n (%)** | 2 (5%) |
| **Rheumatic disease, n (%)** |  |
| RA | 15 (35%) |
| Psoriatic Arthritis | 8 (19%) |
| SLE | 4 (9%) |
| Other Spondyloarthritis | 4 (9%) |
| Sarcoidosis | 2 (5%) |
| Mixed Connective Tissue Disease | 2 (5%) |
| Polymyalgia Rheumatica | 2 (5%) |
| ANCA Vasculitis | 2 (5%) |
| Inflammatory Myositis | 3 (7%) |
| Sjogren's Syndrome | 1 (2%) |
| **Immunomodulation, n (%)** |  |
| TNF Inhibitor* | 11 (26%) |
| Methotrexate Only | 6 (14%) |
| Hydroxychloroquine Only | 5 (12%) |
| Rituximab | 3 (7%) |
| Mycophenolate Mofetil | 1 (2%) |
| Other** | 7 (16%) |
| None | 10 (23%) |
| **Vaccination status, n (%)** |  |
| Not vaccinated | 2 (5%) |
| Vaccinated (Moderna) | 11 (26%) |
| Vaccinated (Pfizer) | 22 (51%) |
| Vaccinated (J&J) | 5 (12%) |
| Missing | 3 (7%) |
| **Time From COVID-19 Infection to PASC Evaluation, days (IQR)** | 239 (202, 327) |

\*Includes TNFi monotherapy and TNFi plus conventional DMARD.
\*\*Includes IL-17 inhibitor (n=2), IL-12/23 inhibitor (n=1), belimumab/hydroxychloroquine (n=1), IL-6 inhibitor (n=1), leflunomide (n=1), and <5mg/d of prednisone (n=1).

### Antigens

The antigens used in the study are as follows: CMV gB (Sino Biological), OC43 Spike/S1/S2 (Sino Biological), HKU1 Spike/S1/S2 (Sino Biological), SARS-CoV-2 D614G Spike/S1/S2/RBD/NTD (Sino Biological), SARS-CoV-2 Nucleocapsid (Sino Biological), VZV gE (abcam), HSV-1 gD (abcam), EBV p18 (Immunetech), Staph aureus - Lukab (IBT bioservices), Tetanus Toxin, Recombinant Heavy Chain Fragment C (Native Antigen), RSV fusion protein (Sino Biological), Measles – Endmonston virus (BioRad Antibodies), Mumps – Enders (BioRad Antibodies), Rubella HPV-77 (BioRad Antibodies), Pneumovax®23 Vaccine (MGH Pharmacy), A/Michigan/45/2015 (H1N1) (Immunetech), B/Phuket/3073/2013 (Immunetech), A/Singapore/INFIMH-16-0019/2016 (Immunetech).

### Antigen-specific isotype titer, FcR-binding, and Avidity

Antigen-specific isotype titer and FcR-binding was measured by a multiplex Luminex, as previously described(*64*). Briefly, carboxylated Magplex microspheres were covalently linked to antigen by ester-NHS linkages using sulfo-NHS (Thermo Fisher) and EDC (Thermo Fisher). Immune complexes were formed by adding antigen-coupled microspheres and appropriately diluted plasma (1:100 for IgG2, IgG3, IgA1 and IgM; 1:500 for IgG1; 1:1000 for FcRs) to 384-well plates. Plates were incubated overnight at 4°C, shaking at 700 rpm. The following day, plates were washed with assay buffer (1x PBS with 0.1% BSA 0.02% Tween-20). Antigen-specific isotype titer was detected through the addition of PE-coupled mouse anti-human detection antibodies (Southern Biotech). To detect antigen-specific FcR-binding, Avi-tagged FcRs (Duke Human Vaccine Institute) were biotinylated with a BirA500 kit (Avidity). The biotinylated FcRs were then fluorescently tagged using streptavidin-PE (Agilent) and added to immune complexes. Fluorescence was read using an iQue (Intellicyt). The data represents the median fluorescence intensity (MFI). The assay was run in duplicate, and the data reported shows the average of the replicates. Each experiment was performed with technical duplicate.

Avidity assays were carried out as antigen-specific duplicate isotyping experiments as described above. One replicate was washed with 7M urea after the immune complex stage while the other was washed with PBS at room temperature. Avidity index was calculated according to the following formula: Avidity index = (Mean MFI sample + Urea)/(Mean MFI sample - Urea).

### Antibody-dependent neutrophil phagocytosis (ADNP)

The antibody-dependent neutrophil phagocytosis (ADNP) assay was performed as previously described(*64*). Fresh peripheral whole blood was collected from healthy donors at the Ragon Institute. The study was approved by the Massachusetts General Hospital Institutional Review Board. Donors were over the age of 18 and provided signed, informed consent. All samples were deidentified prior to use. Red blood cells were lysed using ammonium-chloride-potassium lysis. White blood cells were washed and resuspended at a concentration of 2.5×10^5^ cells/mL in RPMI supplemented with RPMI-1640 (Sigma Aldrich) supplemented with 10% fetal bovine serum (FBS), 5% penicillin/streptomycin (Corning, 50 μg/mL), 5% L-glutamine (Corning, 4 mM), 5% HEPES buffer (pH 7.2) (Corning, 50 mM) for the assay. Biotinylated antigen was coupled to yellow green Neutravidin beads (Invitrogen) and antigen-coupled beads were washed and mixed with diluted plasma (1:50) in 96-well plates to form immune complexes. These plates were incubated for 2 hours at 37°C and washed after the incubation. White blood cells were added to the plates and incubated for 1 hour at 37°C. Neutrophils were stained using mouse anti-human CD66b Pacblue (Biolegend). Fluorescence was acquired using an iQue. The PhagoScore was calculated as follows (GeoMean bead+ cells x %bead+ cells)/10000. The assay was run in duplicate and the data reported represents the average of the replicates.

### CMV Seropositive Analysis

We used the distribution plot of CMV gD IgG1to determine whether individuals who had likely been exposed to CMV in our cohort. We defined samples with log MFI over 4 were defined as seropositive.

### Antibody Depletions

With a four-fold molar excess, CaptureSelect IgM Affinity Matrix (ThermoFisher Scientific) was used to deplete IgM, Protein G Protein Agarose (ThermoFisher Scientific) was used to deplete IgG, and PBS was added to undepleted mock samples overnight at 4C. Isotype-specific depletion was confirmed with a multiplexed Luminex Human isotyping assay (Millipore Sigma) and fluorescence was acquired on an iQue (Intellicyt) flow cytometer.

### Statistical Analysis

Statistical analysis was performed in R (version 4.0.0) or python (version 3.9.1). Prior to analysis, Luminex and ADCD data was log10-transformed and all data was centered and scaled.

For univariate analysis, significance was determined by a two-sided Mann-Whitney U test. For classification models, the systemsseRology R package (v1.0) (https://github.com/LoosC/systemsseRology) was used. Partial least squares discriminant analysis was built using the elastic net-selected features, validated by comparing the cross-validation of the built model with the null permuted labels models.

### Ratio Analysis

To investigate the ratios of antibody features in our two groups with an unsupervised approach we calculated all permutations of antibody feature ratios. To interrogate patterns of OC43 antibody affinity maturation that would result in an increase of one isotype and a corresponding decrease in a second, we investigated the ratios of all measured antibody features for each individual OC43 antigen in Figure 3E, i.e., FcR3a OC43 S2 / IgM OC43 S2. We next calculated the log2 Fold Change (FC) for each ratio between individuals with and without PASC and then tested for statistical significance differences for each ratio between the same groups with Mann-Whitney U testing. We then used a volcano plot of log2 FC and -log2 p-value to display antibody feature ratios that were most enriched in PASC vs. individuals without PASC.

We also used this unsupervised approach to interrogate which antigen specific FcR signals are enriched in PASC individuals. In Figure 4C, we investigated the ratio permutations of OC43 and Spike antigens per FcR binding assay, i.e., OC43 Spike FcR3a / SARS-CoV-2 Spike FcR3a. As described above, we then constructed a volcano plot of log2 FC vs. - log2 P-value to display antigen feature ratios of FcR binding that were most enriched in individuals with PASC vs. without PASC.

## Supporting information

Supplementary Materials for Herman et al. "Impact of cross-coronavirus immunity in post-acute sequelae of COVID-19"

## Data Availability

All data produced in the present study are available upon reasonable request to the authors.

## Acknowledgements

We thank Nancy Zimmerman, Mark and Lisa Schwartz, an anonymous donor (financial support), Terry and Susan Ragon, and the SAMANA Kay MGH Research Scholars award for their support. We acknowledge support from the Ragon Institute of MGH, MIT and Harvard, the Massachusetts Consortium on Pathogen Readiness (MassCPR), the NIH (3R37AI080289-11S1, R01AI146785, U19AI42790-01, U19AI135995-02, U19AI42790-01, 1U01CA260476 – 01, CIVIC75N93019C00052, R01 AR077607, P30 AR070253, P30 AR072577, K23AR073334, 1UL1TR002541-01, and R03AR078938), the Gates Foundation, the Global Health Vaccine Accelerator Platform funding (OPP1146996 and INV-001650), the Musk Foundation, R. Bruce and Joan M. Mickey Research Scholar Fund, the Rheumatoid Research Foundation, and the Doris Duke Charitable Foundation.

J.D.H., Z.W., J.S., and G.A. conceived of the idea. C.E.C., N.J.P., K.M.V, E.N.K., G.Q., N.A.S., Z.W., and J.S. designed, conducted, analyzed the clinical cohort. J.D.H., C.A, and G.A. designed the experiments. J.D.H., C.A., and Y.Z. performed the experiments. J.D.H. and C.A. analyzed the data. J.D.H., C.A., D.L., Z.W., J.S., and G.A. wrote the paper with input from all authors.

G.A. is a founder of SeromYx Systems, Inc. and is a member of the scientific advisory board of Sanofi Pasteur. J.S. has received research support from Bristol Myers Squibb and performed consultancy for AbbVie, Amgen, Boehringer Ingelheim, Bristol Myers Squibb, Gilead, Inova Diagnostics, Janssen, Optum, and Pfizer unrelated to this work. Z.W. reports research support from Bristol-Myers Squibb and Principia/Sanofi and consulting fees from Horizon, Sanofi, Viela Bio, Zenas BioPharma, Shionogi, and MedPace. The other authors declare no competing interests.

The dataset generated during and/or analyzed during the current study have been made available in the Supplementary material.

