## Supplementary Materials for Herman et al. "Impact of cross-coronavirus immunity in post-acute sequelae of COVID-19" for "Impact of cross-coronavirus immunity in post-acute sequelae of COVID-19"

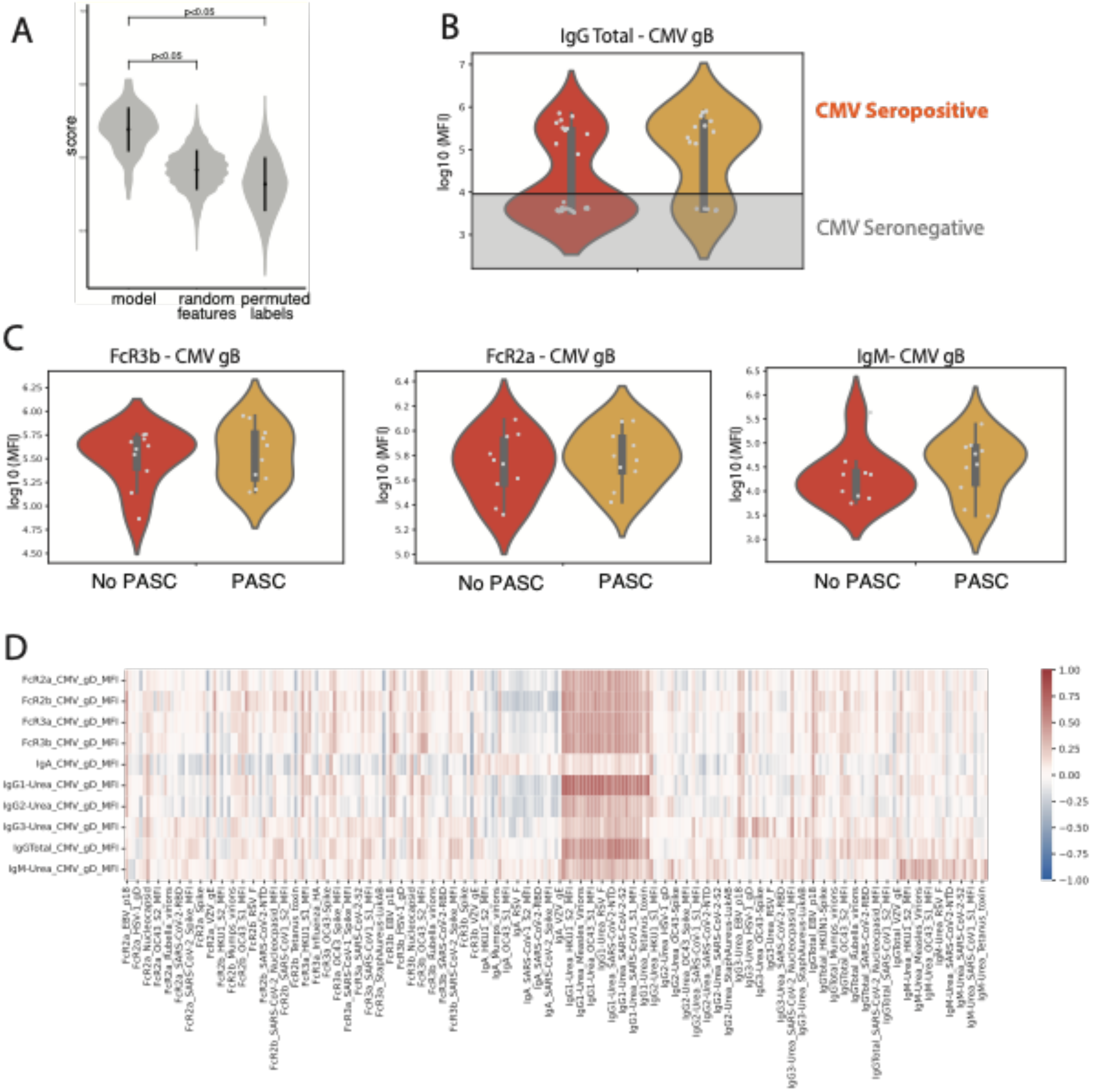


**Figure S1. Supplementary Figure for Figure 2**

(A) Violin plot of 5-fold Cross validation approach for Elastic Net PLS-DA model in Figure 2 A and B. The model was compared to null models with Random features and with Permuted sample labels for 100 replicates. Significance is determined by a two-sided permutation testing and the p-values are indicated in graph.

(B) The violin plot shows CMV gB Total IgG for the PASC and No PASC groups. Samples with log MFI over 4 were defined as seropositive. Significance was determined by a two-sided Mann-Whitney test, * $p = 0.05$, ** $p < 0.05$.

(C) The violin plots show CMV gB TFcR3b, FcR2a, and IgM for CMV seropositive PASC and No PASC groups. Significance was determined by a two-sided Mann-Whitney test, * $p = 0.05$, ** $p < 0.05$.

(D) A heatmap of Spearman correlation coefficients of CMV associated features (Y-axis) with the larger antigen panel including corona virus and other common infectious antigens.

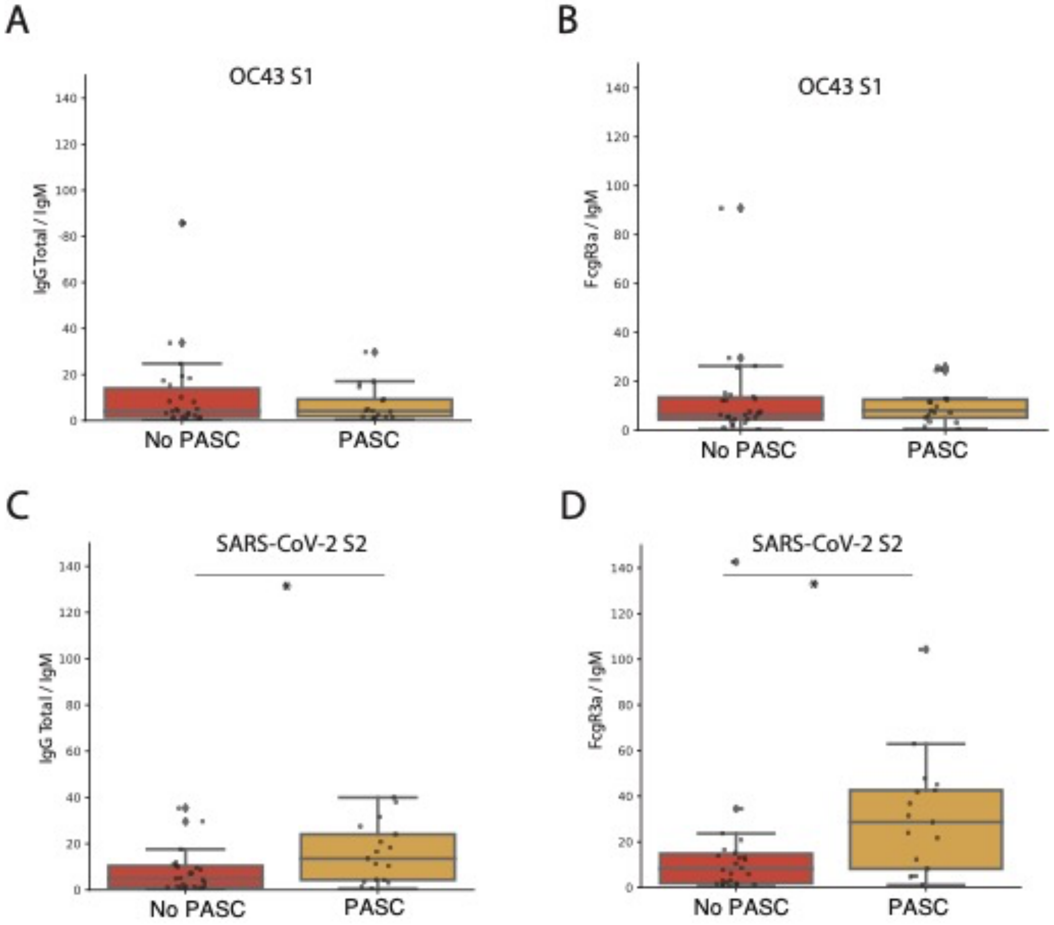


**Figure S2. Supplementary Figure for Figure 3**

Box plots show selected ratios in PASC (Yellow) and No PASC (Red) individuals for (A) the ratio of IgGtotal/ IgM for OC43 S1 domain and (B) of FcR3a/IgM for OC43 S1 domain (C) the ratio of IgGtotal/ IgM for SARS-CoV-2 S2 domain and (B) of FcR3a/IgM for SARS-CoV-2 S2 domain. Significance was determined by a two-sided Mann-Whitney U test. * $p < 0.05$.

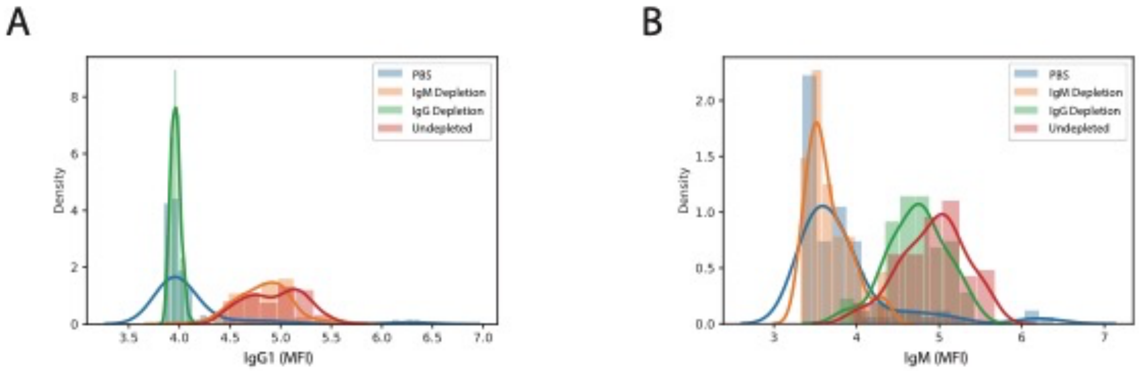


**Figure S3. Supplementary Figure for Figure 4**

(A, B) Confirmation of isotype depletion was performed with Luminex human isotyping Kit. The histograms show the distribution of fluorescence intensities in the Undepleted, IgG depleted, and IgM depleted serum samples. (A) Distribution of IgG1 signal in the three sample sets. (B) Distribution of IgM signal in the three sample sets.
